# AI-Simulated Clinical Consultations: Assessing the Potential of ChatGPT to Support Medical Training

**DOI:** 10.1101/2025.10.07.25337156

**Authors:** Arpita Saggar, Vania Dimitrova, Duygu Sarikaya, David C. Hogg, Jonathan C. Darling

**Affiliations:** School of Computer Science, University of Leeds; Leeds Institute of Medical Education, School of Medicine, University of Leeds

## Abstract

**Background:** Simulated medical scenarios are useful for evaluating and developing clinical competencies but scheduling them is expensive and time-consuming. Large language models (LLMs) show promise in role-playing tasks. We investigated the fidelity with which ChatGPT can mimic patients, clinicians and examiners in educational settings.

**Objective:** To determine the realism with which ChatGPT can portray patient, doctor and examiner roles, and the utility of these agents in clinical education.

**Method:** We selected four paediatric scenarios from mock OSCEs and set up separate patient, doctor and examiner ChatGPT agents for each. The patient and doctor agents conversed with each other in written format. The examiner agent marked the doctor agent based on this conversation. Patients and clinicians familiar with the OSCE assessed the dialogues.

**Results:** The patient agent was judged to be true to character most of the time and good at expressing emotion. The doctor agent was reported to be an effective communicator but occasionally used jargon. Both agents tended to produce repetitive responses which undermined realism. The examiner agent had good correlation with human clinicians. There was moderate support for using the simulated interactions for educational purposes.

**Conclusion:** Although the realism of the agents can be improved, ChatGPT can generate plausible proxies of participants in medical scenarios and could be useful for complementing standardised patient (SP)-based training.

**KEY MESSAGES:** *What is already known on this topic:*

- LLM-based agents show promise for portraying clinical roles and supporting simulation-based learning. Doctor agents provide correct diagnoses most of the time, while patient agents can accurately relay role information such as medical history or symptoms.

*What this study adds:*

- There is scope for improvement in the realism and authenticity of the conversations produced by GPT patient and doctor agents. Notable issues included a tendency to produce repetitive and verbose responses, and an inability to accurately convey the hesitation shown by real patients.
- Disparities observed between (human) patient and clinician assessment for the GPT agents suggest that diverse viewpoints are needed to fully capture the experiential learning associated with clinical communication.
- How this study might affect research, practice or policy
- Low fidelity of GPT simulations for difficult or challenging medical scenarios necessitates human oversight and correction for AI deployed in educational settings.
- The impact of AI on medical education is likely to increase in the future, which necessitates promoting AI literacy among educators and students.

## 1 INTRODUCTION

The development of effective clinical communication skills is vital to ensure the delivery of quality healthcare. Simulated medical scenarios have been widely used in medical education settings for decades. They are a reliable method for teaching these skills, as well as evaluating the competencies of doctors [1], through assessments such as the Objective Structured Clinical Examination, or OSCE [2]. In these simulations, patient roles are typically fulfilled by actors who have been coached to present like real patients, using pre-determined scenarios [3]. However, recruiting and training standardised patients is time-consuming. Scheduling interactions also requires administrators to maintain records and book the SPs. Additionally, the total cost of remuneration to SPs may be substantial for large-scale training and assessment [4,5], which further reduces the feasibility of such arrangements. There is increasing pressure on clinical placement time due to workforce expansion [6].

Recent advances in generative artificial intelligence (AI), especially in large language models, offer hope for cost-effective ways to supplement clinical and communication skills training. These models are tuned to follow instructions, which include adopting a given persona over the course of an interaction [7,8]. They can converse while portraying specific roles, making them ideal for simulation-based teaching and assessment. However, the utility and reliability of these models in the medical classroom remains unclear [9]. Many previous efforts focus on measuring the performance of LLMs on standardised medical examinations [10–14]. While useful for establishing clinical knowledge, these evaluations rely solely on quantitative metrics and fail to explore the capabilities of language models in open-ended discourse. Other studies that leverage LLM-based doctor agents for generating patient-facing dialogue tend to prioritise performance on rigid attributes like information gathering ability, diagnostic reasoning, or compliance to guidelines [15–17]. However, effective clinical communication extends beyond providing accurate diagnoses, and requires interpersonal skills such as building rapport, providing reassurance, and displaying empathy. A notable exception which incorporates this assessment is the AMIE (Articulate Medical Intelligence Explorer) system by Google [18]. AMIE extensively fine-tunes an LLM for clinical conversations rather than using an off-the-shelf model. Studies that simulate patient agents to produce clinician-facing dialogue are less common. Assessment criteria for these largely focus on fidelity to assigned role and the ability to recall information [19–23]. Few have incorporated analyses of dialogue realism and authenticity [15,24]. Furthermore, qualitative evaluations of LLM agents tend to rely solely on medical expertise and fail to take patient perspectives into account. A more comprehensive investigation into the conversational abilities of LLM-based agents is needed before they can be deployed for medical training.

To address this investigative shortfall and better establish the opportunities and challenges of adopting modern LLMs for simulation-based teaching, we conducted a study to evaluate the fidelity of clinical interactions simulated using ChatGPT-based agents. ChatGPT is a popular large language model by OpenAI that has been trained to follow user instructions. We opted for an off-the-shelf model since it is easily accessible through web and programming interfaces. Moreover, fine-tuning billion parameter models requires significant computational resources, which are unlikely to be available in most academic settings. Our aim was to determine the realism with which the model can play the role of patients, doctors and examiners.

## 2 METHODS

We selected four paediatric scenarios typical of the undergraduate OSCE and prompted ChatGPT to simulate a patient agent, a doctor (student) agent and an examiner agent for each scenario. These agents are subsequently referred to as PatientGPT, DoctorGPT and ExaminerGPT. We simulated all three participants to fully explore the breadth of educational applications that could be supported with AI, such as peer-based learning (DoctorGPT) and providing narrative feedback (ExaminerGPT). Using prompts designed from the OSCE station vignettes, PatientGPT and DoctorGPT were instructed to converse with each other. The simulated dialogues were then passed to ExaminerGPT for grading DoctorGPT’s performance. Finally, clinicians and patients familiar with the OSCE assessed the realism of the generated conversations and their utility as a teaching supplement. Clinicians also graded DoctorGPT’s performance, and their marking was compared with that of ExaminerGPT to measure the overlap between human and AI judgement. We used the GPT-3.5 Turbo model^1^ in a zero-shot setting (i.e., without providing any examples of expected outputs) for all experiments. The model was accessed through the OpenAI application programming interface (API) in Python. The scenarios were sourced from mock paediatrics OSCE stations aimed at Year 4 MBChB students within the School of Medicine at the University of Leeds. They were chosen purposefully to reflect the main communication objectives commonly assessed in OSCEs, including information giving, history taking, and managing difficult or sensitive situations. The four stations are:

1. Exploring a mother’s concerns about the MMR^2^ vaccine’s potential link to autism, and discussing its risks and benefits.
2. Explaining test results, and meningitis diagnosis and treatment to a baby’s father.
3. Taking paediatric history and explaining management for a benign heart murmur to a child’s father.
4. Taking an adolescent history from a teenage girl using the HEADSSS ^3^ approach [25] and explaining self-harm management.

For each scenario, we set up one simulation each of PatientGPT, DoctorGPT and ExaminerGPT. Station-specific instructions from the OSCE vignettes were converted to the second person to set roles for the agents. Additional instructions to elicit desirable behaviours were appended to the station-specific instructions. These were identified by iteratively refining the role-setting prompts for each agent. PatientGPT was prompted to use colloquial language, express emotions and relate to life experiences (where appropriate), and use filler words to enhance the realism of its responses. The DoctorGPT agent was instructed to be sensitive and empathetic, provide comprehensive answers, summarise key points at the end of the conversation, and recommend relevant sources of information (where appropriate). No instructions about OSCE time limits were added to the prompts since the model did not have access to any external tools to keep time. The conversations were synthesised by prompting DoctorGPT to initiate a dialogue with PatientGPT. Responses were exchanged between the agents until a response signalling the end of the conversation was produced. The simulated interaction was then provided to ExaminerGPT to grade DoctorGPT’s performance. The grading included station criteria, communication skills, organisation skills and overall grade, all of which were taken from the OSCE information sheets. When no marking style was specified, we found that ExaminerGPT tended to grade very leniently, so we added an additional instruction for strict grading.

Expert evaluation was done by three paediatric consultants and three expert patient volunteers^4^ involved in delivering the OSCE. The number of assessors was chosen consistent with similar recent studies [15,16,24]. The protocol for evaluation was reviewed by the School of Medicine Research Ethics Committee, University of Leeds on 27 January 2024 (Reference: MREC 23-023). Informed consent was sought from all human participants, and all identifiable data was anonymised before analysis. No demographic data were collected. Since the OSCE time limit was not considered during experimentation, the evaluators were asked to disregard conversation length while assessing the outputs. They rated PatientGPT and DoctorGPT on qualitative attributes and provided free-text feedback. They also rated the pedagogical utility of the simulated dialogues. Clinicians graded DoctorGPT’s performance using the same criteria and rubric as ExaminerGPT. All assessments were collected electronically using online forms. Weighted Cohen’s kappa [26] was used to measure the level of agreement between each pair of clinicians, as well as between each clinician and ExaminerGPT. The prompts for agents, the simulated conversations, and the free-text feedback from assessors are provided in the online supplementary material.

## 3 RESULTS

### 3.1 PatientGPT

#### 3.1.1 Qualitative Attributes

PatientGPT was judged on faithfulness to assigned role, the ability to express emotions, and perceived authenticity, using a five-point scale (Figure 1). Results for each assessor group have sample size 12 (3 assessors * 4 stations), while the combined results have sample size 24 ((3+3) assessors * 4 stations). PatientGPT was judged to be moderately faithful to its assigned character (15/24 in agreement overall), with the patient assessment being more favourable than the clinician assessment (9/12 vs 6/12 in agreement respectively). Similar results were obtained for the authenticity of the patient agent (16/24 in agreement overall), with higher approval from the patient group than the clinician group (10/12 vs 6/12 in agreement respectively). Judgement on the emotions expressed in PatientGPT’s responses was more consistent, with both clinicians and patients reporting high agreement (11/12 and 10/12 respectively).

**Figure 1:**
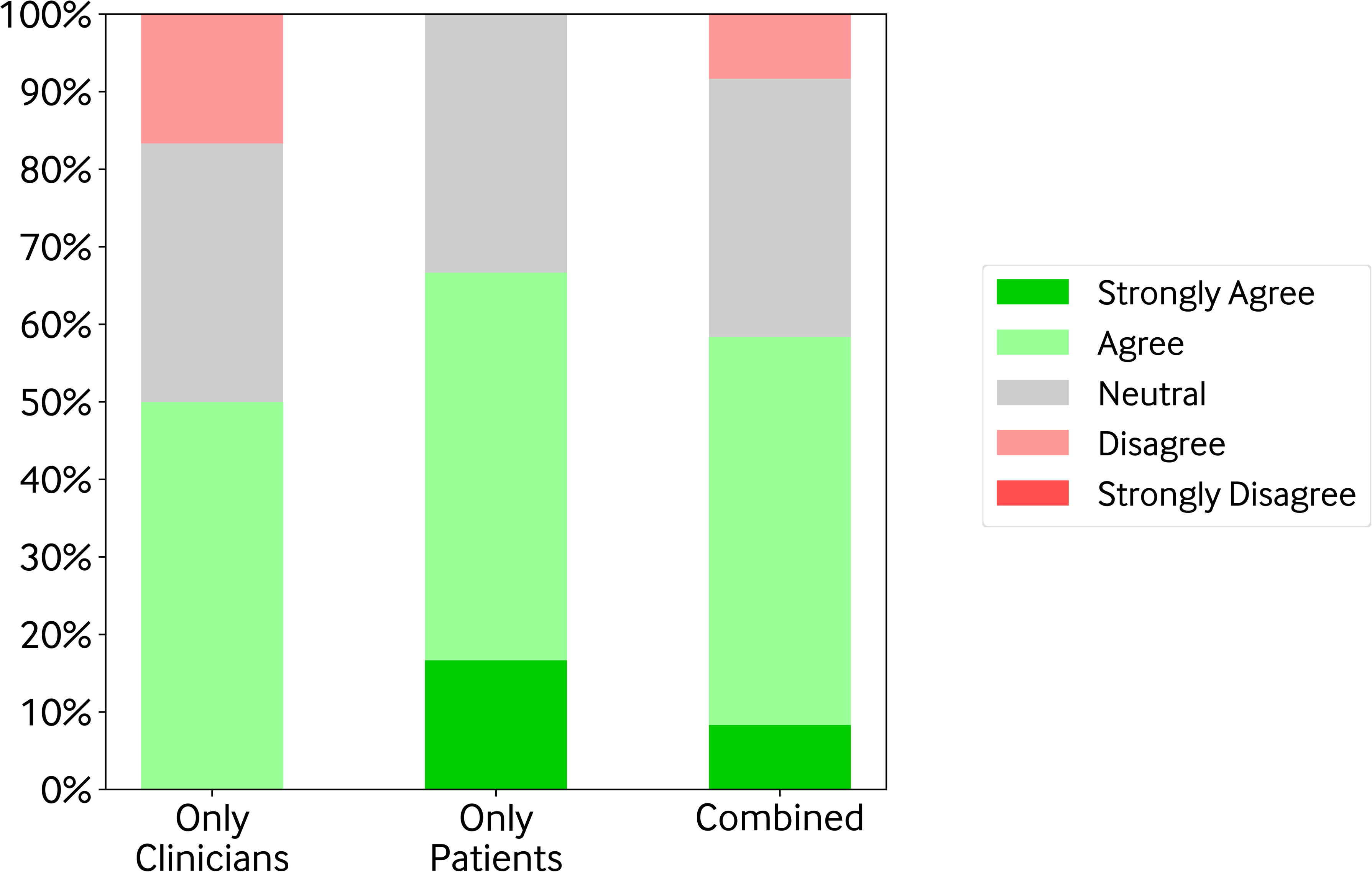
Expert evaluations for the PatientGPT agent across all scenarios. The ’Only Clinicians’ and ’Only Patients’ bars represent a sample size of 12, while the bar for ’Combined’ depicts percentages based on 24 samples.

#### 3.1.2 Free-text feedback

Patients and clinicians also provided open-ended feedback for the LLM agents. Some repetition and unnaturalness were reported for PatientGPT’s responses. An example of this echoing (from the benign heart murmur case) is shown below:

**DoctorGPT:** “…That’s good to hear that Jack has been doing well and hasn’t had any significant symptoms since the viral infection.”

**PatientGPT:** “Yeah, it’s definitely a relief that Jack has been doing well and hasn’t had any more symptoms. I was a bit worried when the GP mentioned the murmur, you know?”

It was also noted that responses by the patient agent were over-eager and did not accurately mirror the hesitation shown by actual patients. This was especially relevant in the self-harm management scenario (17 sensitive content below):

**DoctorGPT:** “…Can you tell me a bit more about what happened and how you’re feeling now?”

**PatientGPT:** “Yeah, sure. So, earlier tonight, I got into this big argument with my boyfriend. It was, like, really intense and we ended up breaking up. I was so upset and I just couldn’t see a life without him, you know? So, I did something stupid and cut my wrists with a razor blade. But now, I regret it and I don’t want to die anymore. I just want to forget this whole thing happened.”

The clinician group additionally commented that unlike actual patients, PatientGPT was persuaded more easily to change its stance and opinions. This behaviour may be attributed to the wider issue of *sycophancy* in LLMs [27], where models trained on human feedback tend to tailor responses by repeating or reinforcing the user’s preferences (the user in this scenario being DoctorGPT). Other issues included occasional repetition of DoctorGPT’s jargon and overuse of colloquial expressions which diminished the perceived concern. However, both groups reported that PatientGPT’s responses were appropriate most of the time.

### 3.2 DoctorGPT

#### 3.2.1 Qualitative Attributes

Qualitative assessment for DoctorGPT was based on communication skills, organisation skills and the ability to show empathy. Figure 2 reports the ratings received by the agent, along with the corresponding rating scales. The ‘patient only’ and ‘clinician only’ results have sample size 12 (3 assessors * 4 stations), while the combined results have sample size 24 ((3+3) assessors * 4 stations). DoctorGPT was reported to be an empathetic communicator overall (20/24 in agreement), with the patient ratings being slightly more varied across the five-point scale than those from the clinician group (10/12 in agreement for both groups). Both groups favoured the second highest rating (on a four-point scale) for the agent’s communication skills, with the patient group tending to award slightly higher scores than the clinicians. Perceptions on the organisational approach of DoctorGPT were less consistent. The patient group leaned towards the highest rating, while ratings from the clinician group were almost equally divided between ‘Average’ and ‘Good’ scores.

**Figure 2:**
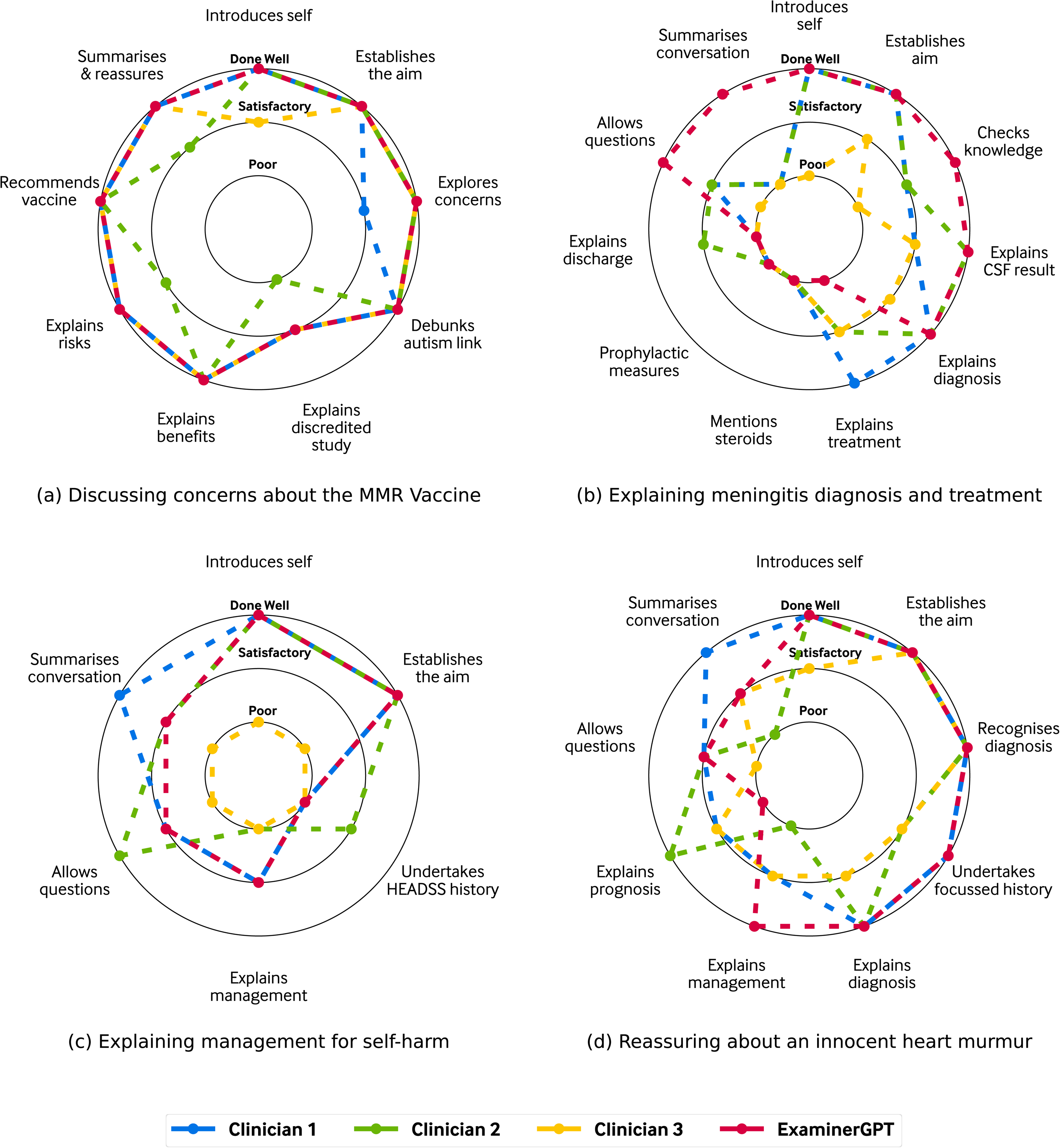
Evaluation for DoctorGPT’s empathy, communication skills and organisational skills. The ’Only Clinicians’ and ’Only Patients’ bars represent a sample size of 12, while the bar for ’Combined’ shows percentages corresponding to 24 samples.

#### 3.2.2 Station Grades

Clinicians also assessed station-specific criteria and provided an overall grade for the doctor agents (Table 1) using a five-point scale (Clear fail, Borderline, Clear Pass, Very good pass, Excellent Pass). The rubric corresponding to the grades is provided in the <u>online supplementary material</u>. DoctorGPT received passing grades from all assessors in the MMR vaccine and heart murmur stations. Assessment for the meningitis diagnosis scenario was also in favour of (borderline) passing grades. There was no overlap in clinician marking for the self-harm scenario, but the (human) majority was again in favour of (different) passing grades.

**Table 1:** The overall grades awarded to DoctorGPT for each OSCE station.

| **Station** | **Clinician 1** | **Clinician 2** | **Clinician 3** | **ExaminerGPT** |
|---|---|---|---|---|
| **MMR Vaccine** | Very good pass | Very good pass | Clear pass | Clear pass |
| **Meningitis Diagnosis** | Borderline | Borderline | Clear fail | Borderline |
| **Self-harm management** | Borderline | Clear pass | Clear fail | Clear fail |
| **Heart Murmur** | Clear pass | Clear pass | Clear pass | Borderline |

#### 3.2.3 Free-text feedback

Recurring themes in DoctorGPT’s feedback included wordy, repetitive answers which undermined realism, and lack of summarisation of key points of the conversation. The agent also failed to recommend resources for further information where needed. Some responses were reported to be formulaic and lacking personalisation. The patient group additionally reported that DoctorGPT sometimes followed a tick-box approach, focussing on follow-up questions rather than letting PatientGPT elaborate concerns, which was deemed somewhat unnatural. However, DoctorGPT was judged by both assessor groups to be good at providing reassurance and explanations, despite occasionally responding with unnecessary jargon. This was observed in the MMR vaccine scenario, where the agent included journal names and exact statistics when debunking the link between the vaccine and autism:

**DoctorGPT:** “…One notable study published in the Journal of the American Medical Association (JAMA) in 2002 included over 500,000 children and found no increased risk of autism following the MMR vaccine. Another study published in the New England Journal of Medicine in 2019 analyzed data from over 650,000 children and also found no association between the MMR vaccine and autism…”

Additionally, while DoctorGPT received a (borderline) passing grade for the self-harm management case, it failed to mention drug use or sexual relationships, which are vital aspects of the HEADSSS history. This may be an artefact of continuing efforts to detoxify outputs by restricting topics of conversation [28].

### 3.3 ExaminerGPT

Performance of ExaminerGPT was measured by checking the overlap between its markings and human judgement. Both clinicians and ExaminerGPT marked DoctorGPT on station-specific criteria using a three-point scale (Done well, Satisfactory, Not attempted or Poor). Figure 3 shows how the four evaluators marked DoctorGPT for each station. Across the four scenarios, the average agreement between ExaminerGPT-Clinician pairs (κ = 0.54,0.36,0.26; mean = 0.39) was higher than the average agreement between Clinician-Clinician pairs (κ = 0.42,0.22,0.35; mean = 0.33). For overall grade (Table 1), ExaminerGPT’s judgement had reasonable overlap with the clinicians for binary pass/fail decisions, but overlap with the majority judgement occurred in only one case (Meningitis Diagnosis). The justifications produced by ExaminerGPT for its scoring are available in the <u>online</u> <u>supplementary material</u>.

**Figure 3:**
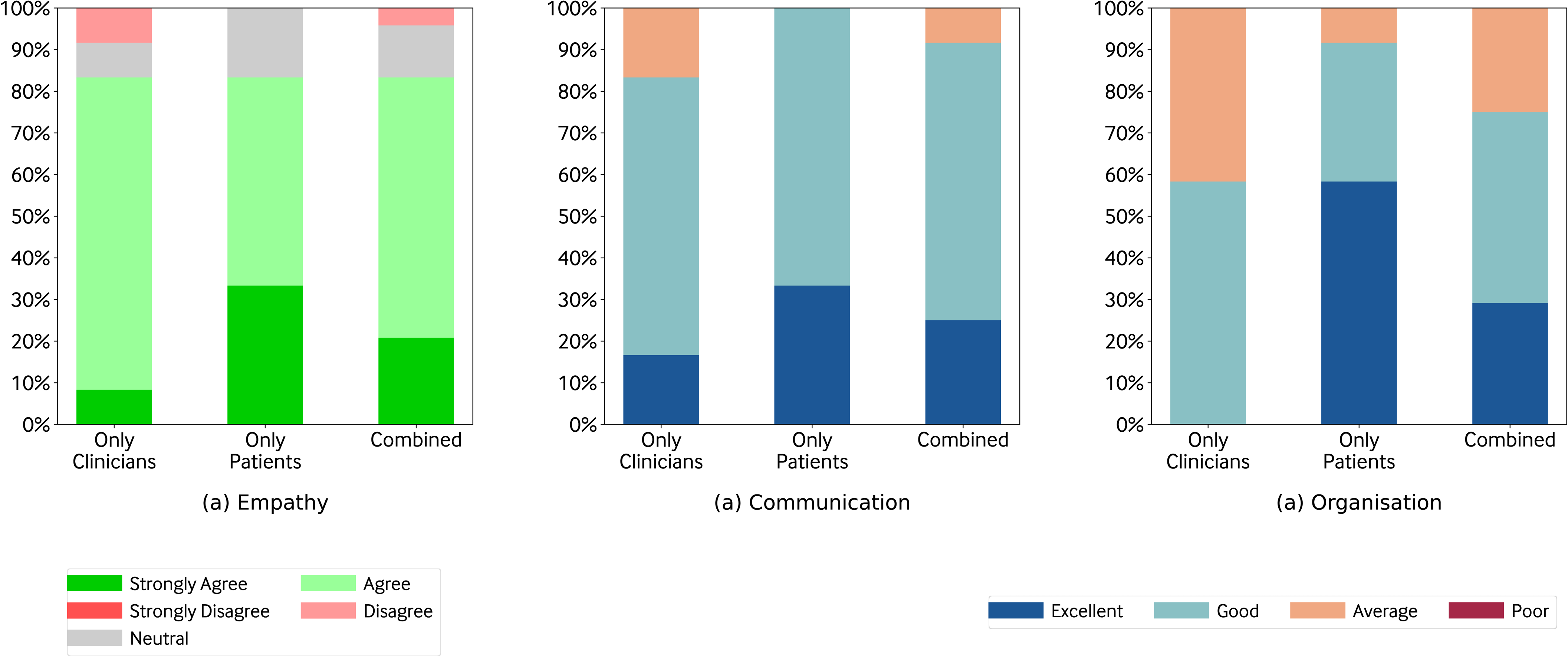
Station-specific marking for DoctorGPT done by three human clinicians and ExaminerGPT. The agent had good overlap with clinician judgement most of the time, but struggled in cases where a stricter assessment was requisite.

### 3.4 Pedagogical Utility

The final question posed to both patients and clinicians addressed the potential utility of the simulated interactions in medical training. Figure 4 presents assessor agreement with the statement: “*This conversation can be used for educational purposes*”. Just over half of the combined assessors thought the conversations could be a useful learning resource (14/24 in agreement), with the patient response being more favourable than the clinician response (8/12 vs 6/12 in agreement).

**Figure 4:**
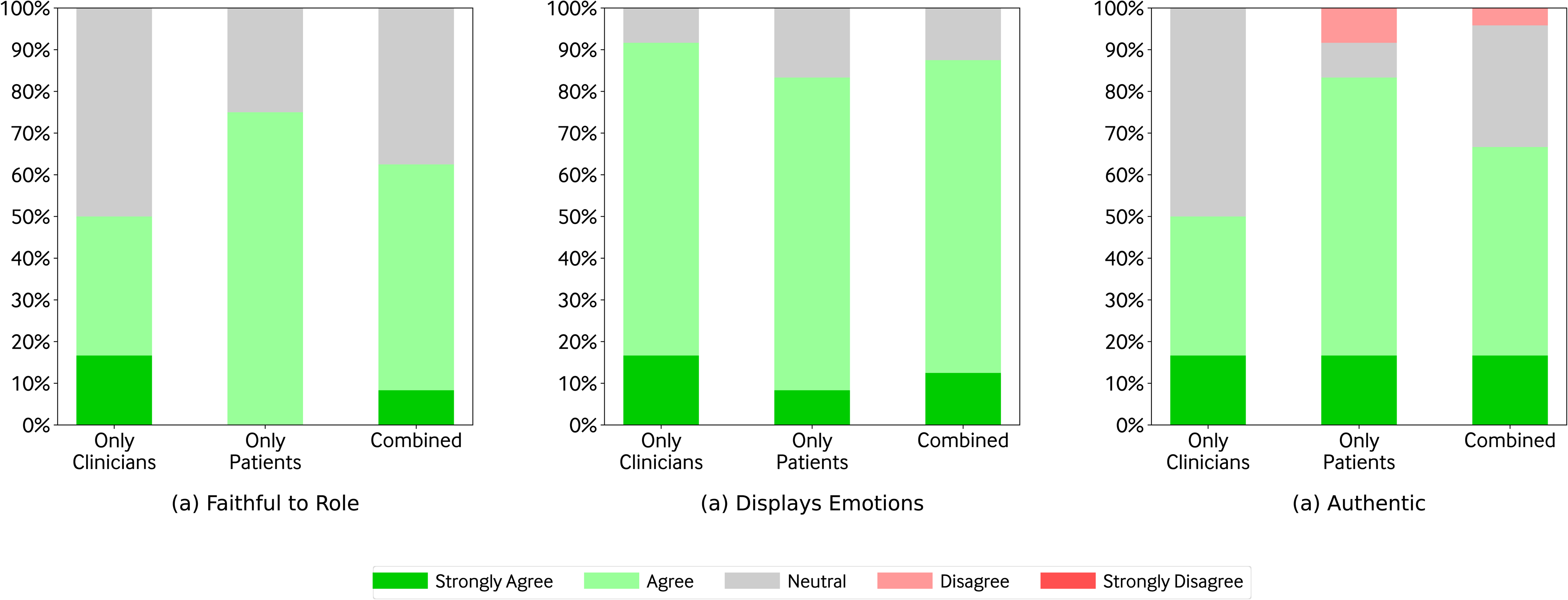
Assessor responses on the usefulness of the simulated dialogues as a learning aid. The ’Only Clinicians’ and ’Only Patients’ bars represent a sample size of 12, while the bar for ’Combined’ shows percentages corresponding to 24 samples.

## 4 DISCUSSION

Overall, we found reasonable evidence of realism offered by the ChatGPT-based simulated agents. PatientGPT showed good ability to express emotions while responding. The agent was faithful to its character for most of the interaction and moderately representative of true patients. DoctorGPT was good at expressing empathy and offering explanations, but failed to address some sensitive aspects of clinical communication. The agent also had scope for improvement in its organisational skills. However, DoctorGPT attained passing grades in all scenarios (based on majority clinician ratings). ExaminerGPT displayed high agreement with the human clinicians, but tended to mark leniently. There was moderate support for using such simulated conversations in clinical teaching.

Our findings largely corroborate recent studies that showcase the potential of large language models in supporting simulation-based training [19,24]. However, many works have restricted assessment of agents to rigid measures such as diagnostic correctness [15–17] or the ability to recall role-specific information [20,22,23]. In contrast, we have sought qualitative assessments as well as free-text feedback, which provide greater insight into the behaviour of the GPT agents. A notable issue which undermined the perceived realism of both agents was a tendency to frequently agree with and repeat each other’s statements, resulting in some unnatural echoing and verbosity. As a result, both assessor groups reported that PatientGPT was more amenable to changing its opinion than real patients. Although this *sycophancy* can limit the applicability of GPT-based agents for simulating diverse scenarios with angry or difficult patients, we found limited discussion on this issue in the existing literature [24]. Learning to personalise communication based on patient characteristics is also essential for experiential learning [29]. The patient group reported that DoctorGPT tended to favour formulaic approaches to conversation. Despite being a potential barrier to building trust and rapport with patients, few studies have considered this facet in their assessment [18]. Finally, while existing literature relies solely on medical expertise to evaluate LLM-based agents, the current study also incorporates expert patient perspectives. Notable disparities between assessments done by the two assessor groups were observed for PatientGPT’s faithfulness to role and authenticity, and for DoctorGPT’s organisational skills. This was despite both assessor groups being familiar with the OSCE, and highlights the subjectivity inherent in discourse evaluation, as well as the importance of including diverse perspectives.

While the role-playing capabilities of ChatGPT are limited for complex scenarios, it can still serve as a valuable learning tool, particularly for formative assessment and feedback. Reviewing representative conversational transcripts can help learners improve their interviewing skills [30]. These dialogues can also be used to finetune smaller language models for role-playing, which can then be deployed in resource-constrained settings. Conversely, scenarios scored poorly by assessors could be used to help learners identify and reflect on missed opportunities. Large language models offer a scalable method to educate students about consultation skills in the clinical workplace, encompassing both common and rare scenarios. However, there are intrinsic limits to the consistency and reliability of their outputs [31], which necessitate a continuing need for human oversight of models deployed in pedagogical settings. Our study also highlights the growing need to train faculty in AI-enhanced teaching, covering prompt engineering and tool integration. Finally, we note some limitations in our work. Our findings are likely to have limited generalisability in diverse cultural settings due to the small number of scenarios simulated, and a relatively small assessor group. They are also limited to the written format and do not incorporate non-verbal aspects of communication such as tone of voice or body language. Furthermore, we have not considered the OSCE time limit in our experiments, and results will likely differ when this aspect is incorporated. Future studies may explore incorporation of real-world constraints, and novel prompting strategies to reduce sycophancy and repetition in GPT-generated dialogues.

## Data Availability

All data produced are available online at https://arpita2512.github.io/ai_simulated_clinical_consultations/

## ACKNOWLEDGEMENTS

The authors are grateful to the clinicians and patients who took part in the evaluation, and the Patient and Carer Community at the Leeds Institute of Medical Education, University of Leeds. We have also benefitted from helpful comments provided by Dr. Matt Homer on a draft of this work.

## Footnotes

1 June 13, 2023 snapshot with 4k context window

2 Measles, Mumps and Rubella

3 Home environment, Education & Employment, Activities, Drugs, Sexuality, Suicide/Depression, Safety from injury and violence

4 Members of the Patient and Carer Community at the Leeds Institute of Medical Education

